# Effects of age, period, and birth cohort on fall-related mortality in older adults in Brazil from 1980 to 2019

**DOI:** 10.1101/2024.07.19.24310703

**Authors:** José Mário Nunes da Silva, Rita de Cássia de Lima Idalino

**Affiliations:** Department of Statistics, Federal University of Piauí, Teresina, Brazil; Laboratório de Inferência Causal em Epidemiologia (LINCE – USP), University of São Paulo, São Paulo, Brazil; Laboratório de Colaboração Estatística (LCE – UFPI), Federal University of Piauí, Teresina, Brazil

**Keywords:** falls, mortality, age effect, period effect, cohort effect, older people

## Abstract

**Aim:** This study aimed to estimate the effects of age, period, and birth cohort on fall-related mortality in older adults in Brazil and its geographic regions, by sex, from 1980 to 2019.

**Methods:** We conducted an ecological time-series study using data on fall-related deaths in older people extracted from the official mortality information system. Poisson models were adjusted for each sex and geographic region to estimate age-period-cohort effects.

**Results:** From 1980 to 2019, Brazil recorded 170,607 fall-related deaths in older adults, with 50.1% occurring in women. More than half of these deaths occurred in the 80 years or older age group (55.0%) and in the Southeast region (52.0%). We observed an increase in fall-related mortality rates (FMR) across all age groups and regions, regardless of sex. There was an increased risk of death in all periods after the reference period (2000 to 2004) in all geographic regions and for both sexes. We also observed a gradual increase in mortality risk for men born before 1914 and after 1935 compared to the reference cohort (1930 to 1944). In contrast, we found a protective effect across all birth cohorts for women.

**Conclusion:** There was a consistent increase in fall-related mortality risk among older people in Brazil, posing a public health challenge. The findings highlight the urgent need for implementing public health policies promoting older adult health and preventing fall risks to improve the quality of life for this population.

**What is new?:** *Key Findings:*

- The study found a consistent increase in fall-related mortality rates (FMR) among older adults in Brazil from 1980 to 2019.
- Age Effect: Fall-related mortality rates (FMR) increased progressively with advancing age. More than half of the fall-related deaths occurred in individuals aged 80 years or older (55.0%).
- Period Effect: There was an increased risk of death in all periods after the reference period (2000 to 2004) across all geographic regions and for both sexes.
- Cohort Effect: For men, there was a gradual increase in mortality risk for those born before 1914 and after 1935 compared to the reference cohort (1930 to 1944). For women, a protective effect was observed across all birth cohorts.

*What This Adds to What Was Known?:*

- This study is one of the first to analyze fall-related mortality trends in Brazil using the age-period-cohort (APC) model.
- Traditional studies have typically focused on time trends of standardized rates, which consider age and period effects but often overlook the impact of different birth cohorts.

*What Is the Implication and What Should Change Now?:*

- The findings highlight the urgent need for public health policies to promote older adult health and prevent falls, focusing on both healthcare and physical environments to mitigate increasing mortality risk.
- Enhancing public health strategies to prevent falls can improve the quality of life for the elderly population in Brazil and mitigate the public health challenge posed by the rising trend in fall-related mortality.

## Introduction

Population aging is a global reality that brings significant changes to the epidemiological profile of populations^1^. In Brazil, increased life expectancy and declining birth rates have led to a rapid growth in the proportion of older people^2^. This natural process causes various physiological and systemic changes in older adults, which can compromise balance and increase the risk of falls^3,4^.

The World Health Organization (WHO) defines a fall as an event which results in a person coming to rest inadvertently on the ground, floor, or other lower level^5^. Globally, approximately 684,000 individuals die from falls annually^5^, leading to nearly 17 million years of life lost^6^. In Brazil, 15.5% of individuals over 60 experience falls annually, rising to 22.3% after age 75^7^. Those who have fallen before face a 60% to 70% risk of recurrence within a year, with a 20% mortality rate^8^. Furthermore, healthcare costs for older persons fall victims rise annually^9–11^.

The literature extensively describes the problem of falls among older adults^6,10,12,13^. However, few or no studies have been conducted exploring trends in fall-related mortality rates (FMR), considering the effects of age, period, and birth cohort (APC). Traditional studies often restrict their analysis to examining time trends of standardized rates^10,11,14–19^, which assess age and period effects but may miss how different generations are affected by evolving health practices and socioeconomic conditions over their lifespans^20^.

APC models allow for decomposing these effects, providing valuable insights into the specific influence of each factor, representing an advancement in the trend analyses normally used in epidemiology. Therefore, understanding these dimensions is crucial for resource allocation and the development of more effective and targeted public health interventions aimed at reducing the incidence of falls and their fatal consequences among older people. We aim to estimate, for the first time, the effects of age, period, and birth cohort on fall-related mortality among older adults in Brazil and its geographic regions, by sex, from 1980 to 2019.

## Methods

### Study Design

We conducted an ecological study examining the temporal trends in fall-related mortality among older people individuals in Brazil and its geographical regions, encompassing both sexes aged 60 years or older, spanning from 1980 to 2019.

### Data sources and population

Mortality data were extracted from the Mortality Information System (SIM), coordinated by the Brazilian Health Informatics Department (Datasus), available at: http://tabnet.datasus.gov.br/. Mortality information included codes E880 to E888 from the International Statistical Classification of Diseases and Related Health Problems, 9th Revision (ICD-9), and codes W00 to W19 from the 10th Revision (ICD-10).

Population data were also sourced from Datasus based on the censuses of 1980, 1991, 2000, and 2010. Population projections for the inter-census years’ July 1st populations were estimated by the Brazilian Institute of Geography and Statistics (IBGE). We adjusted and corrected the mortality data through proportional redistribution of initially classified records with unknown age or sex across each geographical region, study year, sex, and age group before applying the filter for individuals aged 60 years and older in the analyzed data^21^.

### Variables

After obtaining death records and population data, crude and specific annual mortality rates from falls per 100,000 inhabitants were calculated by age group, according to sex and geographic regions, as well as proportional mortality. Subsequently, rates were standardized by age and sex using the direct method, based on the World Standard Population proposed by Segi in 1960^22^.

To improve data stability, ages were grouped into five-year intervals, starting from 60-64 years and ending with 80 years or older, totaling five age groups. Periods were also grouped into five-year intervals, with the first period covering 1980-1984 and the last period covering 2015-2019, totaling eight periods. Finally, birth cohorts ranged from 1900 to 1959, totaling 12 cohorts.

### Statistical analysis

The effects of age, period, and birth cohort were calculated assuming a Poisson distribution for fall-related deaths, with temporal effects (age-period and birth cohort) acting multiplicatively on the rate. Therefore, the logarithm of the expected rate is a linear function of the effects of age, period, and cohort, as observed in Equation 1 below^23^:

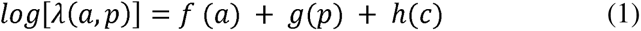

Where *λ*(α*, p*) represents the mortality rate at age *α* and period *p* for individuals in birth cohort *c* = *p*-*α*; *f*, *g*, *h* are functions of age, period, and cohort, respectively, which can be parametric or non-parametric; *α*, *p* and *c* denote the average age, period, and cohort for the observational units^23^.

The primary challenge in adjusting a model involving age, period, and cohort is their linear relationship. For instance, when age and period are known, cohort can be inferred. This limitation is known as the "non-identifiability problem"^24^, and there is no consensus on the best solution. In our study, we opted to estimate the parameters of the APC effect using Carstensen’s proposed method^23^. This method often involves parameterizing age, period, and cohort effects in a way that allows for their separation, often using techniques such as splines to smoothly model variables while avoiding imposing rigid structures. Thus, models with different forms for functions *f*, *g*, *h* were adjusted: a factor model with one parameter per level, and two models using smoothing functions, natural splines, and B-splines^23^. Ultimately, we selected the function that provided the best fit.

We compared models using likelihood ratio tests to assess the effects of age, period, and cohort. Submodels were systematically adjusted to identify the nonlinear effects of these explanatory variables: age, age-drift (cohort drift model), age-cohort, age-period-cohort, age-drift (period drift model). Based on these comparisons, the best model was obtained. We verified the adequacy of the final model fit using deviance statistics^23^.

Based on estimates from the final model, we graphically expressed period and cohort effects as relative risks (RR) of fall-related mortality compared to a reference period and cohort, respectively. We used the reference cohort from 1930 to 1935 and the period from 2000 to 2004, chosen because central cohorts and periods typically exhibit greater stability^25^. We presented age effects graphically as fall-related mortality rates (per 100,000 inhabitants) by age, adjusted for the reference cohort and period. We considered values statistically significant through analysis of 95% confidence intervals. The APC model estimation analyses were conducted using the “Epi” package in the open-source software R version 4.3.3 (R Foundation for Statistical Computing, Vienna, Austria).

## Results

Between 1980 and 2019, Brazil recorded 170,607 deaths from falls among older adults, with 50.1% occurring in women. More than half of these deaths were observed in the age group of 80 years or older (55.0%) and in the Southeast region (52.0%). When comparing the years 1980 and 2019, an increase in crude FMR was observed across all age groups and regions, as well as in standardized FMR for both men and women (Table 1). The highest standardized FMR for men (40.8/100,000) and women (28.5/100,000) were observed in 2018 and 2017, respectively. The Central-West region exhibited standardized FMR 2.5 times higher than the region with the lowest rates, the Northeast. It is noteworthy that all regions showed a progressive increase in standardized FMR over the study period, with the highest rates evident after 1998 (Figure S1).

**Table 1.** Fall-related mortality rates among older adults (per 100,000 inhabitants), by age, sex, and geographic regions. Brazil, 1980-2019.

| Age groups (years) | Men |  |  |  | Women |  |  |  |
| --- | --- | --- | --- | --- | --- | --- | --- | --- |
|  | N | % | Rate |  | N | % | Rate |  |
|  |  |  | 1980 | 2019 |  |  | 1980 | 2019 |
| North |  |  |  |  |  |  |  |  |
| 60-64 | 417 | 14.0 | 2.4 | 14.2 | 138 | 5.2 | 5.1 | 2,6 |
| 65-69 | 430 | 14.4 | 10.5 | 11.8 | 195 | 7.4 | 8.4 | 10,5 |
| 70-74 | 422 | 14.2 | 8.9 | 20.6 | 276 | 10.1 | 12.8 | 15,8 |
| 75-79 | 454 | 15.2 | 16.7 | 44.7 | 393 | 14.8 | 40.5 | 42,9 |
| ≥80 | 1.259 | 42.2 | 166.2 | 156.0 | 1.656 | 62.5 | 10.5 | 154,3 |
| Standardized rate |  |  | 22.0 | 30.4 |  |  | 11.1 | 24.6 |
| Northeast |  |  |  |  |  |  |  |  |
| 60-64 | 1.671 | 11.6 | 0.3 | 13.9 | 813 | 5.1 | 1.7 | 4,7 |
| 65-69 | 1.694 | 11.8 | 1.0 | 18.0 | 1.028 | 6.4 | 1.3 | 7,7 |
| 70-74 | 1.833 | 12.7 | 2.4 | 26.4 | 1.555 | 9.7 | 5.9 | 18,3 |
| 75-79 | 2.187 | 15.2 | 3.8 | 43.1 | 2.242 | 14.1 | 4.8 | 36,7 |
| ≥80 | 6.992 | 48.6 | 21.9 | 140.3 | 10.318 | 64.7 | 7.4 | 140,2 |
| Standardized rate |  |  | 3.1 | 31.4 |  |  | 3.2 | 23.2 |
| Southeast |  |  |  |  |  |  |  |  |
| 60-64 | 5.814 | 12.6 | 2.7 | 17.1 | 2.439 | 5.7 | 6.4 | 4,2 |
| 65-69 | 5.873 | 12.7 | 7.9 | 22.9 | 3.006 | 7.0 | 8.4 | 8,9 |
| 70-74 | 6.387 | 13.8 | 23.6 | 31.5 | 4.371 | 10.2 | 15.2 | 16,4 |
| 75-79 | 7.492 | 16.2 | 59.3 | 53.2 | 6.492 | 15.2 | 33.7 | 33,9 |
| ≥80 | 20.572 | 44.6 | 284.2 | 161.4 | 26.336 | 61.8 | 99.5 | 135,9 |
| Standardized rate |  |  | 38.7 | 37.7 |  |  | 19.5 | 22.4 |

| **South** |  |  |  |  |  |  |  |  |
| --- | --- | --- | --- | --- | --- | --- | --- | --- |
| 60-64 | 1.616 | 10.7 | 0.0 | 13.3 | 598 | 3.5 | 1.9 | 5,3 |
| 65-69 | 1.656 | 11.0 | 0.0 | 22.8 | 926 | 5.4 | 4.9 | 9,2 |
| 70-74 | 2.009 | 13.3 | 3.1 | 39.8 | 1.567 | 9.2 | 3.8 | 26,4 |
| 75-79 | 2.502 | 16.6 | 8.8 | 88.3 | 2.454 | 14.4 | 13.3 | 54,1 |
| ≥80 | 7.290 | 48.4 | 335 | 240.0 | 11.510 | 67.5 | 13.5 | 270,5 |
| Standardized rate |  |  | 4.4 | 48.1 |  |  | 5.2 | 38.7 |
| **Central-West** |  |  |  |  |  |  |  |  |
| 60-64 | 702 | 10.6 | 1.6 | 19.0 | 312 | 4.4 | 3.7 | 6,6 |
| 65-69 | 805 | 12.2 | 0.0 | 25.2 | 398 | 5.6 | 2.3 | 9,1 |
| 70-74 | 857 | 12.9 | 7.0 | 43.1 | 606 | 8.5 | 11.5 | 26,5 |
| 75-79 | 975 | 14.7 | 24.3 | 81.4 | 1.119 | 15.7 | 43.4 | 84,2 |
| ≥80 | 3.282 | 49.6 | 109.4 | 287.4 | 4.677 | 65.8 | 67.9 | 306,2 |
| Standardized rate |  |  | 14.0 | 55.1 |  |  | 14.2 | 45.2 |
| **Brazil** |  |  |  |  |  |  |  |  |
| 60-64 | 10.220 | 12.0 | 1.5 | 15.7 | 4.300 | 5.0 | 4.2 | 4,6 |
| 65-69 | 10.458 | 12.3 | 4.2 | 21.2 | 5.553 | 6.5 | 5.4 | 8,7 |
| 70-74 | 11.508 | 13.5 | 12.2 | 31.8 | 8.366 | 9.8 | 10.4 | 19,2 |
| 75-79 | 13.610 | 16.0 | 29.6 | 57.9 | 12.700 | 14.9 | 21.9 | 41,4 |
| ≥80 | 39.395 | 46.2 | 147.0 | 176.0 | 54.497 | 63.8 | 55.0 | 168,0 |
| Standardized rate |  |  | 20.0 | 38.6 |  |  | 11.9 | 26.6 |

Regarding mortality, we observed higher FMR across all age groups after the period between 1995 and 2000, for both sexes, in Brazil and across all geographical regions (Figure 1). For birth cohorts, we found that men belonging to older cohorts experienced a decrease in FMR in the age groups of 75 to 80 years in Brazil and across all geographical regions. For younger birth cohorts, we observed a growing increase in FMR across all age groups in Brazil and all geographical regions, with consistently higher values for male cohorts compared to female cohorts (Figure 2). Additional information is available in the supplementary material.

**Figure 1.**
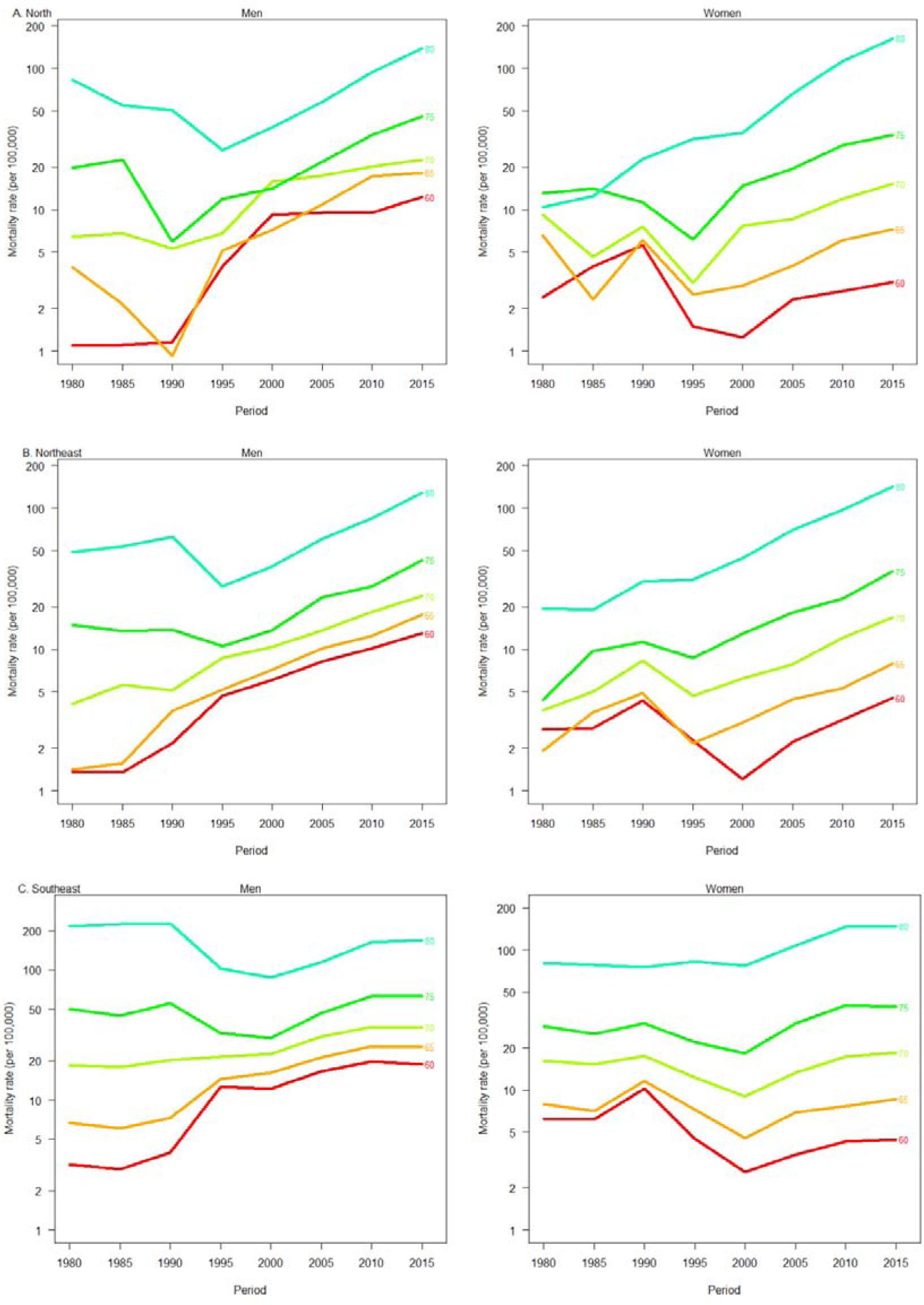
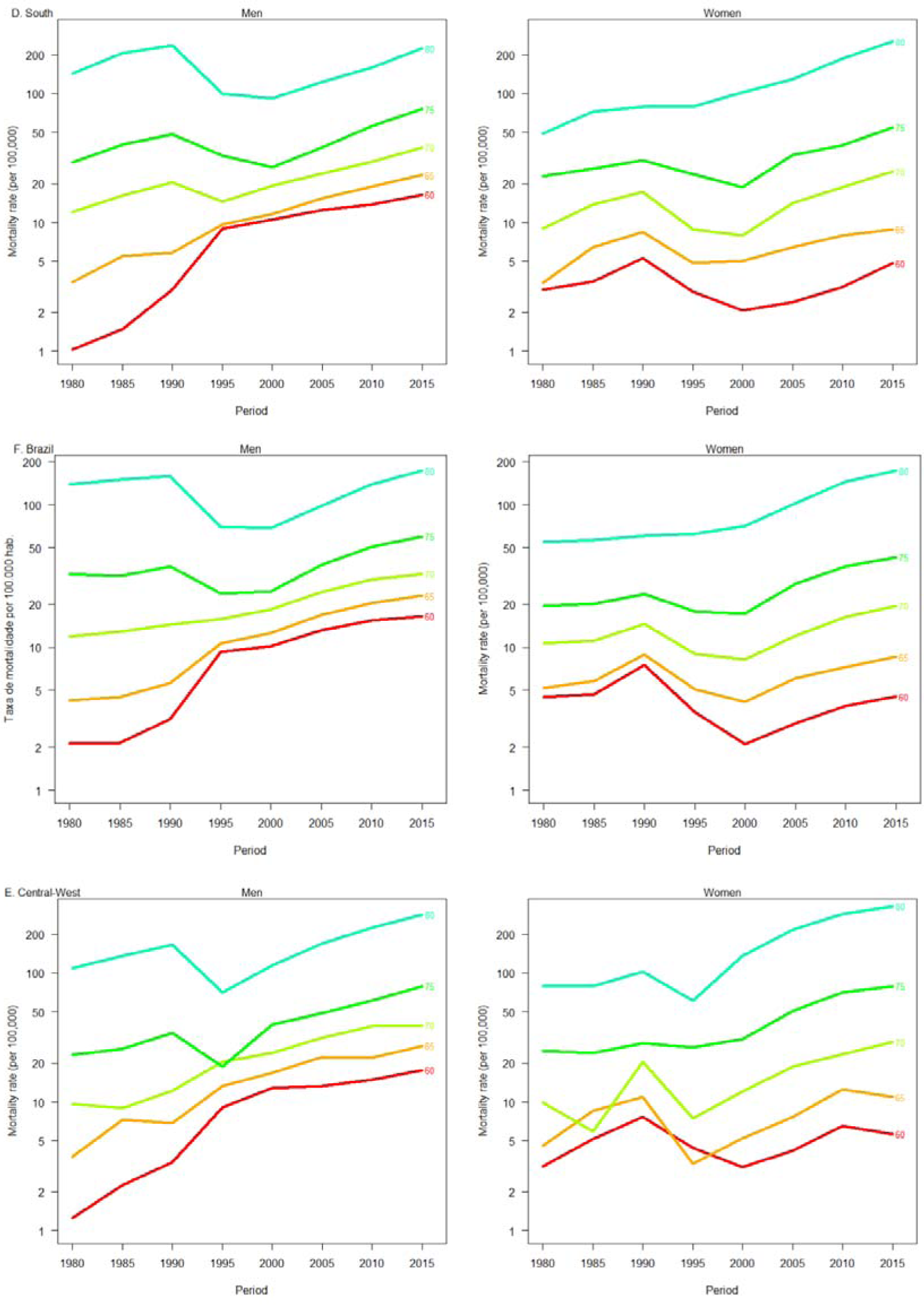
Fall-related mortality rates among older adults per period, connected within each age group, by sex and geographic regions, Brazil, 1980-2019.

**Figure 2.**
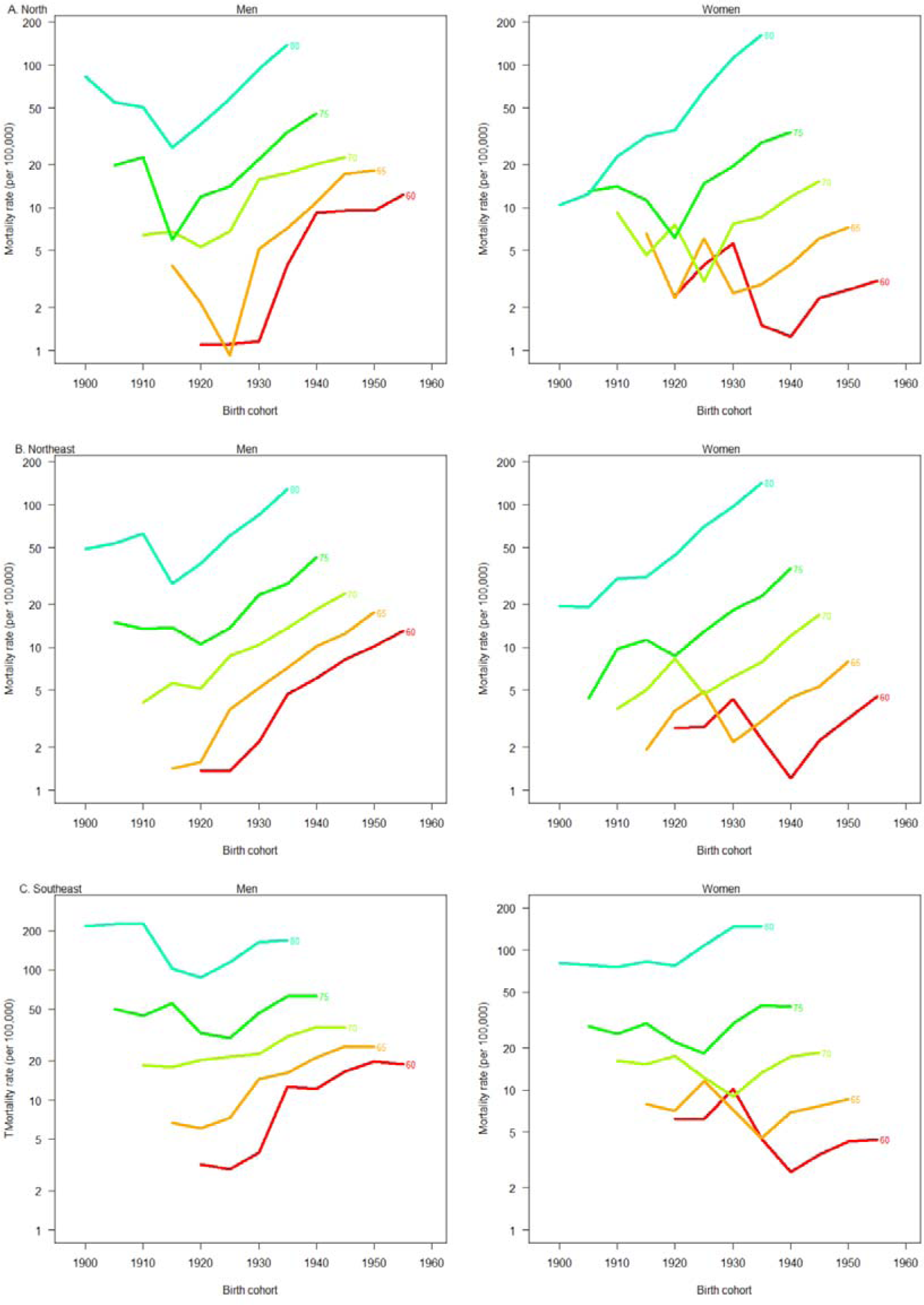
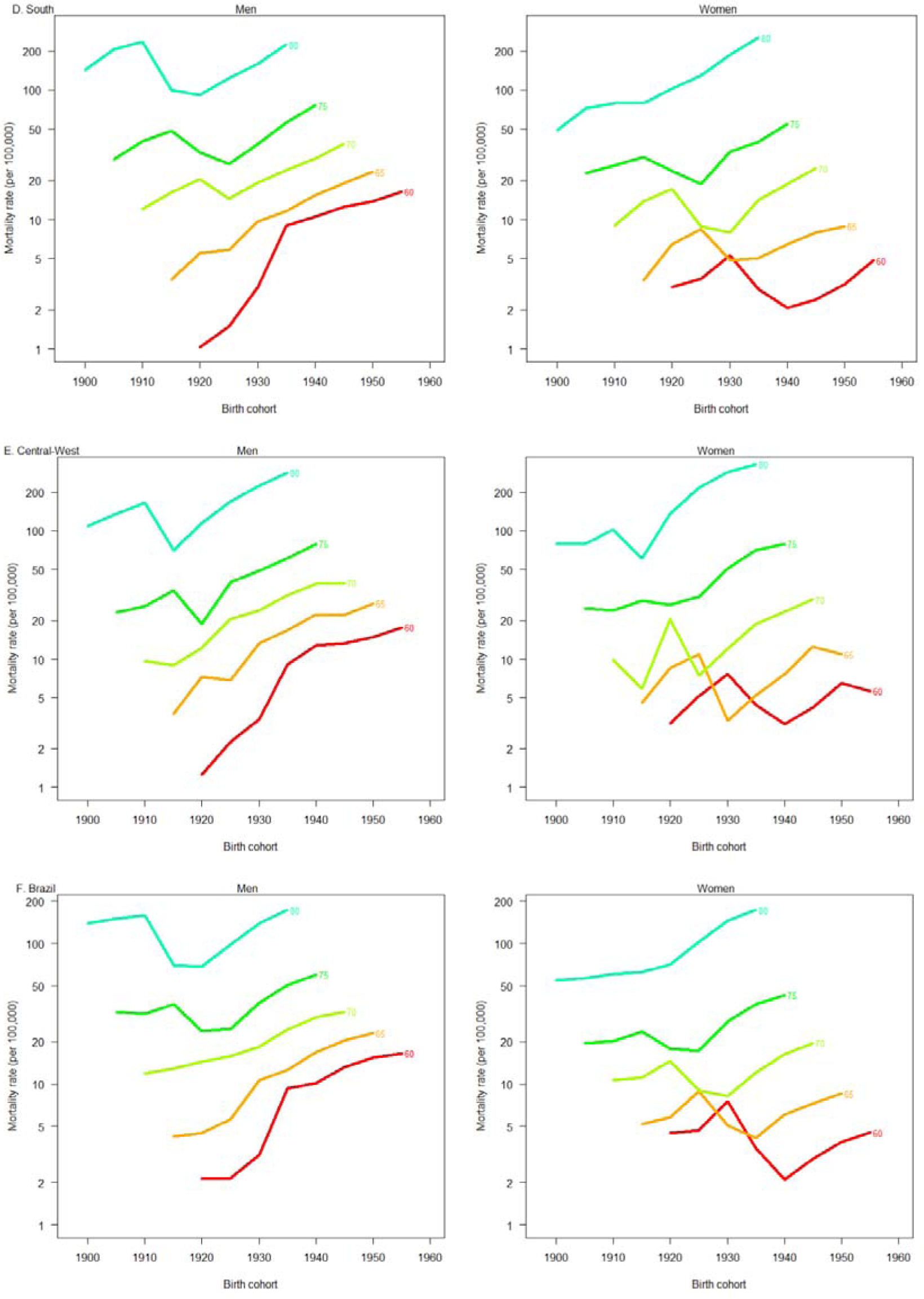
Fall-related mortality rates among older adults by birth cohort, connected within each age group, by sex and geographic regions. Brazil, 1980-2019.

In relation to the analysis of age (A), period (P), and birth cohort (C) effects for both sexes in Brazil and across all geographical regions, we found that the model that best fit the data, with significantly lower deviance (p < 0.0001), was the full APC model (Table 2).

**Table 2.** Model’s estimates for the age-period-cohort effect on fall-related mortality among older adults, by sex and geographic regions, Brazil, 1980-2019.

| Model<sup>a</sup> | Men |  |  |  |  | Women |  |  |  |  |
| --- | --- | --- | --- | --- | --- | --- | --- | --- | --- | --- |
|  | Deviance | df | Deviance<sup>b</sup> | df<sup>b</sup> | p<sup>c</sup> | Deviance | df | Deviance<sup>b</sup> | df<sup>b</sup> | p<sup>c</sup> |
|  | North |  |  |  |  |  |  |  |  |  |
| Age | 827.4 | 35 |  |  |  | 927.7 | 35 |  |  |  |
| Age-drift | 229.2 | 34 | 598.2 | 1 | <0.001 | 251.2 | 34 | 676.5 | 1 | <0.001 |
| Age-cohort | 62.4 | 24 | 166.8 | 10 | <0.001 | 165.9 | 24 | 85.3 | 10 | <0.001 |
| Age-period-cohort | 44.0 | 18 | 18.4 | 6 | <0.001 | 28.7 | 18 | 137.2 | 6 | <0.001 |
| Age-period | 155.1 | 28 | 111.2 | 10 | <0.001 | 171.6 | 28 | 142.9 | 10 | <0.001 |
| Age-drift | 229.2 | 34 | 74.0 | 6 | <0.001 | 251.2 | 34 | 79.6 | 6 | <0.001 |
| Northeast |  |  |  |  |  |  |  |  |  |  |
| Age | 3.859.5 | 35 |  |  |  | 4.891.0 | 35 |  |  |  |
| Age-drift | 850.0 | 34 | 3.009 | 1 | <0.001 | 654.2 | 34 | 4.327 | 1 | <0.001 |
| Age-cohort | 109.3 | 24 | 740.7 | 10 | <0.001 | 543.0 | 24 | 111 | 10 | <0.001 |
| Age-period-cohort | 83.1 | 18 | 26.2 | 6 | <0.001 | 54.4 | 18 | 489 | 6 | <0.001 |
| Age-period | 509.1 | 28 | 426.0 | 10 | <0.001 | 386.5 | 28 | 332 | 10 | <0.001 |
| Age-drift | 850.0 | 34 | 340.8 | 6 | <0.001 | 654.2 | 34 | 268 | 6 | <0.001 |
| **Southeast** |  |  |  |  |  |  |  |  |  |  |
| Age | 5.379 | 35 |  |  |  | 3.189.6 | 35 |  |  |  |
| Age-drift | 4.449 | 34 | 930 | 1 | <0.001 | 1.939.2 | 34 | 1.250 | 1 | <0.001 |
| Age-cohort | 950 | 24 | 3.449 | 10 | <0.001 | 1.470.9 | 24 | 468 | 10 | <0.001 |
| Age-period-cohort | 381 | 18 | 569 | 6 | <0.001 | 216.1 | 18 | 1.255 | 6 | <0.001 |
| Age-period | 3.500 | 28 | 3.119 | 10 | <0.001 | 915.4 | 28 | 699 | 10 | <0.001 |
| Age-drift | 4.449 | 34 | 949 | 6 | <0.001 | 1.939.2 | 34 | 1.024 | 6 | <0.001 |
| **South** |  |  |  |  |  |  |  |  |  |  |
| Age | 2.257.2 | 35 |  |  |  | 3.051.2 | 35 |  |  |  |
| Age-drift | 1.376.3 | 34 | 880.9 | 1 | <0.001 | 658.8 | 34 | 2.392.7 | 1 | <0.001 |
| Age-cohort | 297.1 | 24 | 1.079.2 | 10 | <0.001 | 508.8 | 24 | 149.7 | 10 | <0.001 |
| Age-period-cohort | 140.6 | 18 | 156.4 | 6 | <0.001 | 63.8 | 18 | 445.0 | 6 | <0.001 |
| Age-period | 878.2 | 28 | 737.6 | 10 | <0.001 | 312.8 | 28 | 249.1 | 10 | <0.001 |
| Age-drift | 1.376.3 | 34 | 498.1 | 6 | <0.001 | 658.5 | 34 | 345.6 | 6 | <0.001 |
| **Central-West** |  |  |  |  |  |  |  |  |  |  |
| Age | 1.108.1 | 35 |  |  |  | 1.382.8 | 35 |  |  |  |
| Age-drift | 211.1 | 34 | 897.1 | 1 | <0.001 | 268.3 | 34 | 1.114.5 | 1 | <0.001 |
| Age-cohort | 61.9 | 24 | 149.1 | 10 | <0.001 | 165.5 | 24 | 102.8 | 10 | <0.001 |
| Age-period-cohort | 42.8 | 18 | 19.2 | 6 | <0.001 | 20.6 | 18 | 144.9 | 6 | <0.001 |
| Age-period | 175.0 | 28 | 132.2 | 10 | <0.001 | 126.2 | 28 | 105.6 | 10 | <0.001 |
| Age-drift | 211.0 | 34 | 36.0 | 6 | <0.001 | 268.3 | 34 | 142.1 | 6 | <0.001 |
| **Brazil** |  |  |  |  |  |  |  |  |  |  |
| Age | 12.268.1 | 35 |  |  |  | 12.132.3 | 35 |  |  |  |
| Age-drift<sup>a</sup> | 6.723.0 | 34 | 5.545.1 | 1 | <0.001 | 3.708.7 | 34 | 8.423.6 | 1 | <0.001 |
| Age-cohort | 1.106.8 | 24 | 5.616.3 | 10 | <0.001 | 2.859.7 | 24 | 849.0 | 10 | <0.001 |
| Age-period-cohort | 648.1 | 18 | 458.7 | 6 | <0.001 | 281.1 | 18 | 2.578.6 | 6 | <0.001 |
| Age-period | 4.831.4 | 28 | 4.183.3 | 10 | <0.001 | 1.823.4 | 28 | 1.542.3 | 10 | <0.001 |
| Age-drift<sup>**</sup> | 6.723.0 | 34 | 1.891.6 | 6 | <0.001 | 3.708.7 | 34 | 1.885.3 | 6 | <0.001 |

*Note:* <sup>a</sup>The linear trend of the logarithm of age-specific rates over time equals the sum of the slopes of the period and cohort effects ( $\beta_L + \gamma_L$ ), where $\beta_L$ and $\gamma_L$ are the linear trends for the period and cohort, respectively. <sup>\*\*</sup>The longitudinal age trend is represented by the sum of the linear trends of age and period, where $\alpha$ is the linear trend associated with age and $\beta$ is the linear trend associated with the period
<sup>a</sup>The models are ordered so that adjacent rows provide tests between the models, culminating in the age-period-cohort model.
<sup>b</sup>Changes in residual degrees of freedom and deviance between the models in the current row and the previous row in the table.
<sup>c</sup>P-value of the likelihood ratio test comparing the models in the current row and the previous row in the table.

When considering the effect of age adjusted for period and birth cohort, we observed a progressive increase in FMR with advancing age across all geographical regions of Brazil, regardless of sex. Notably, the Central-West, South, and Southeast regions showed above-average growth compared to the national average and other geographical regions for both sexes (Figure 3 and Table S1).

**Figure 3.**
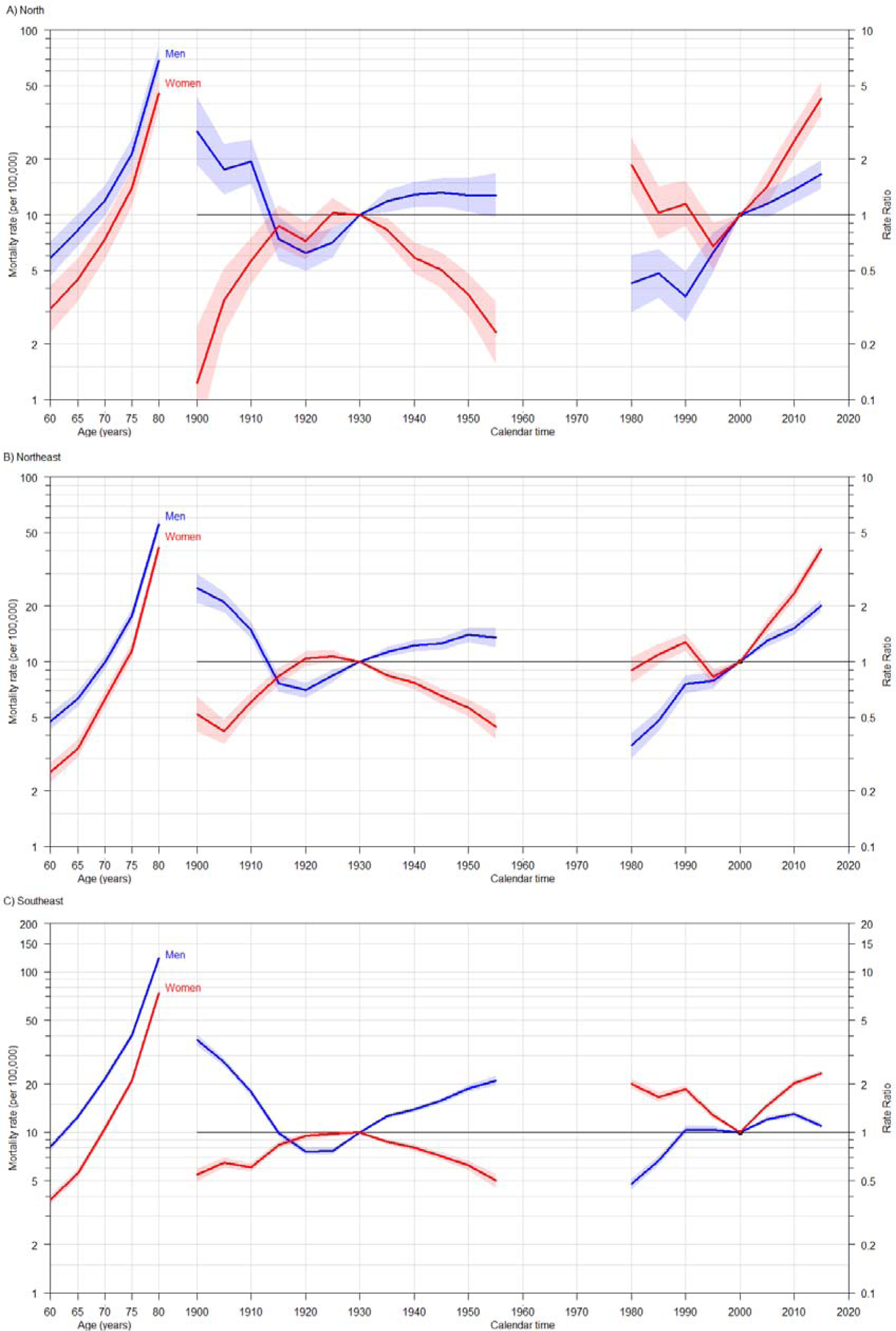
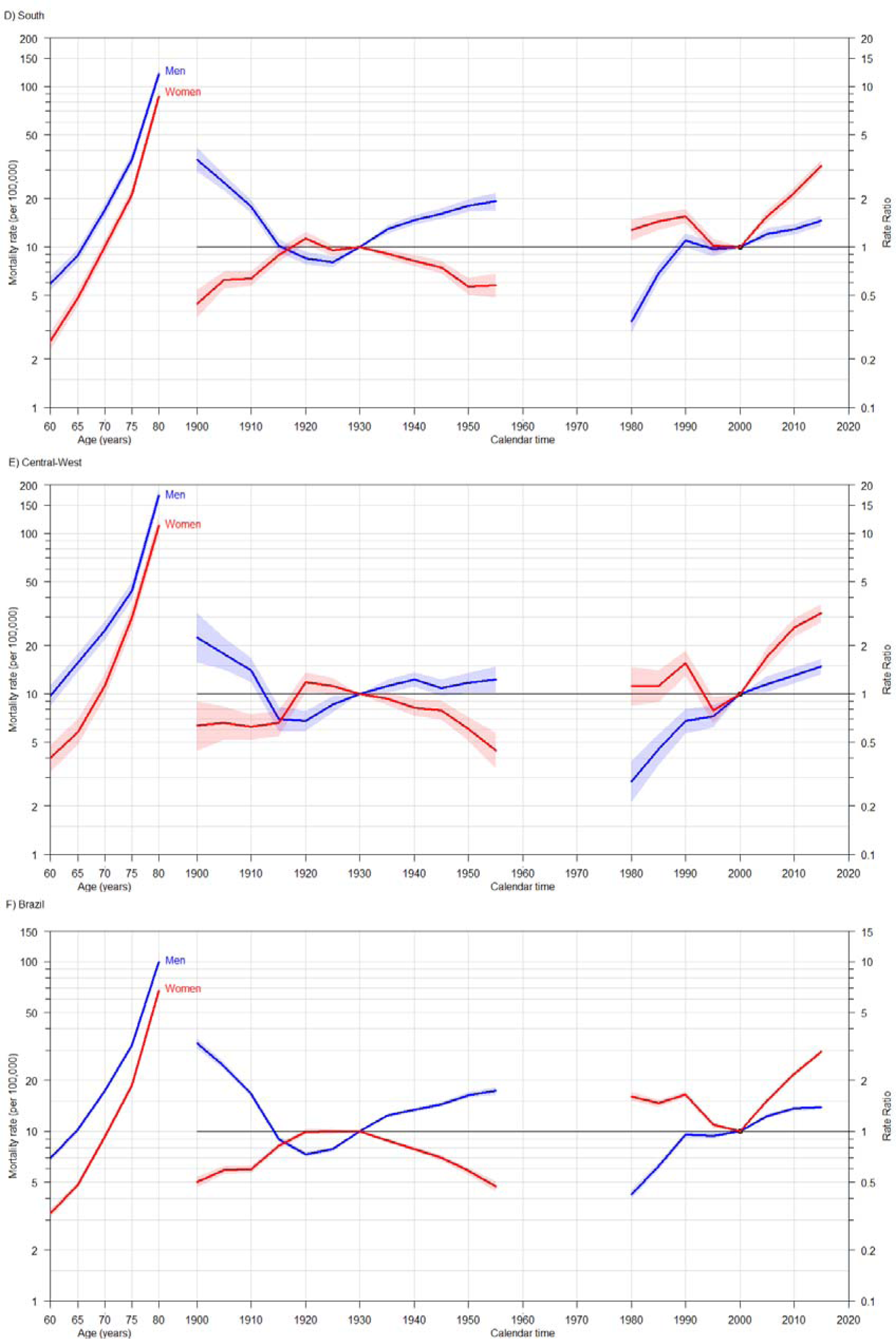
Estimates for age-period-cohort model adjusted for fall-related mortality among older adults, by sex and geographic regions. Brazil, 1980-2019.

Regarding the period effect adjusted for age and birth cohort, we observed a gradual increase in the risk of death from falls in all periods following the reference period (2000 to 2004) across all geographical regions and for both sexes (Figure 3, Table S1, and Table S2).

In terms of the birth cohort effect, after adjusting for period and age effects, we observed a gradual increase in the risk of death from falls for men born before 1914 and after 1935, and a decrease in the risk of death from falls for those born from 1915 to 1929, compared to the reference cohort (1930 to 1944), across Brazil and all geographical regions. Overall, for women, we observed a protective effect across all birth cohorts compared to the reference cohort (Figure 3 and Tables S1 and S2).

## Discussion

This was the first study to analyze the age-period-cohort effect on fall-related mortality among older adults in Brazil, covering deaths over a 40-year period. Our results highlighted age as the most significant and prominent factor for both sexes across all models analyzed. They also revealed higher FMR in older men, particularly in the Central-West, South, and Southeast regions of the country. The period effect emerged as robust and consistent, showing a significant increase from 2000-2004, especially among women in all geographical regions of Brazil. Notably, there were elevated mortality risks among women in the North and Northeast regions during the most recent period, from 2015-2019. Additionally, the cohort effect was significant, with a progressive increase in mortality risk among younger cohorts of men born after 1935, returning to levels similar to those of earlier cohorts before 1915, particularly in the Southeast and South regions, and across Brazil. For women, there was a general reduction in risk across birth cohorts, with the exception of a slight increase in the South and Central-West regions for the 1920-1924 cohorts.

The findings of this study, which show an increase in fall-related mortality among older people with advancing age, are consistent with those of other studies in the literature^6,10,14,15,18,26,27^. This trend can be explained by several interrelated factors. Among these, the increased incidence of falls in this population is notable^28^, influenced by the decline of the musculoskeletal and sensory systems common with aging^29^, as well as sedentary lifestyles, which exacerbate this scenario^3^. The growing prevalence of senile frailty also plays a significant role^17^, linking advanced age and comorbidities to higher fall-related mortality^13,30^. Furthermore, improvements in the quality of mortality records may have contributed to this finding^15^.

It is crucial to emphasize that aging itself is not a direct cause of falls, although the physiological changes associated with it can increase vulnerability^14^. Among older adults, falls usually result from a complex interaction between pre-existing medical conditions and unfavorable environments, which may include physical obstacles or a lack of safety adaptations^3^. These combined factors heighten the risk of falls, which in turn can significantly contribute to the increase in fall-related deaths due to resulting injuries^14,18,30^. This underscores the critical importance of preventive strategies that address both the health conditions and the physical environment of older adults^8^. In our study, FMR showed an increasing trend in the 75-79 age group and rose sharply in those aged 80 or older, highlighting the need for special attention to the health of older individuals. With the progressive aging of the population, it is foreseeable that these numbers will rise in the future^18^. Moreover, the increased frailty and severity of falls in individuals aged 75 and above may explain this trend^17^. Consequently, it would be advisable to implement public health programs both in the community and healthcare settings that prioritize the prevention and/or mitigation of frailty, especially in those aged 75 or older. Randomized trials and systematic reviews have consistently shown that interventions incorporating resistance or strength training and balance exercises for older adults can reduce the risk of both fatal and non-fatal falls^31^, as well as the fear of falling^32^.

While the literature suggests women have a higher risk of falls^3,33^ due to factors like menopause-related decreases in estrogen and vitamin D levels^28,34^, our study did not necessarily reflect this in higher mortality for women. Similar results have been observed in other studies on the topic^14,17,18,35,36^. Notably, across all geographic regions, FMRs were consistently higher among men compared to women, likely due to the circumstances surrounding the falls^33^.

Several authors suggest that higher FMR in men may be due to a higher incidence of outdoor falls, faster leg muscle deterioration, and engagement in more intense activities leading to severe injuries^14,18,37^. In addition, men have more comorbidities and worse health compared to women, exacerbating the consequences of fractures^13,17^. Overcoming the cultural belief that illness is a sign of weakness is crucial to addressing this issue^14^.

Existing literature indicates an increasing trend in fall-related deaths in both high-income and low-to-middle-income countries, across both sexes, similar to our study findings. A recent study conducted in the USA, using trend analysis, found that between 1999 and 2020, there was an annual increase of 3.94% in FMR among adults aged 65 or older^19^. In Spain, from 2000 to 2015, there was an observed annual increase in fall-related mortality trends among adults over 65 years old of 2.5%, with the highest rate found in those aged 75-84 years (6.4%)^17^. In Canada, from 2001 to 2007, falls accounted for 26% of all unintentional injury deaths, being the leading cause of death in women and the second in men^38^. High FMR in adults aged 70 and older was also noted across various European countries between 1990 and 2017^39^. A study in China from 2013 to 2020 estimated an annual increase of 5.08% in FMR among adults aged 60 or older^18^. Similar results have also been observed in trend analyses from other Brazilian studies across different periods and geographic regions analyzed^10,11,14–16,26,27,35^.

Our study found varying mortality risks from falls across birth cohorts and sexes. The decrease in FMR up to 1930 for both men and women may be linked to improvements in health patterns and working conditions during those decades. Advancements in healthcare, including new treatments, medications, and vaccination campaigns, contributed to increased life expectancy and better public health^40^. The establishment of Brazil’s National Department of Public Health in 1920 marked significant progress in public health policies, leading to expanded and more effective health interventions that likely improved quality of life and reduced frailty among older adults^41^. Thus, the protective effect observed in birth cohorts up to 1930 reflects the benefits of these historical improvements in Brazilian public health.

Furthermore, it is crucial to consider the public policies implemented in Brazil after 1935, such as the creation of Labor Laws and the establishment of the Brazilian Unified Health System (SUS)^41^. Specific policies for older people, such as the Active Aging Policy, the National Health Policy for the Elderly, and Statute of the Elderly, had a significant impact on the functionality of this population^4,42^. These improvements allowed, particularly for women, a reduction in the risk of death in younger cohorts. In contrast, the increased risk of death among men in younger cohorts can be attributed to factors such as nutritional transitions, reduced physical activity, exposure to more hazardous working conditions, and the rise in chronic diseases predisposing to falls^18^. We also observed geographic disparities in FMR among older adults in Brazil. Sociodemographic factors may contribute to higher FMR in economically developed regions like the Southeast and South, where urbanization is more prevalent and potentially increases urban obstacles that raise the risk of falls^17^. The North and Northeast regions have younger age structures compared to other regions, potentially resulting in lower incidence of fatal falls among older people^3,13^. Additionally, the cooler climate in the Southeast and South may lead to less physical activity among older adults, increasing muscular weakness and the likelihood of severe falls that lead to mortality^18,38^.

The study has several limitations. Firstly, being an ecological study, the associations observed at the population level may not reflect individual-level outcomes. Secondly, we relied on secondary data for our analyses, so the quality and completeness of mortality information may vary across the study period and between different regions, influencing FMR estimates. Finally, analyzing the age-period-cohort effect is complex and subject to specific assumptions. Therefore, future studies could benefit from including additional variables and employing more sophisticated analysis methods to further explore the results.

Despite these limitations, this study represents the first attempt using an APC model to assess the trend of fall-related mortality over the past four decades in Brazil, disaggregating data by geographic region and sex. This approach is particularly significant as most existing studies only consider the effects of age and period of death, neglecting the impact of birth cohorts on fall mortality trends. Consequently, we observed an increased risk of fall-related deaths among older adults attributed to the effects of age and period in both sexes and across all Brazilian regions. Regarding the effect of birth cohort, we noted an increased risk only in younger cohorts of men across all geographic regions.

## Data Availability

The relevant data are available in the manuscript and supplementary material. Data that not presented are available from the corresponding author upon reasonable request.

## Conclusion

Given these findings, it is evident that there is an urgent need to enhance accident and fall prevention strategies for older people in Brazil. It is imperative that these efforts encompass education, professional training, the creation of safer environments, and interventions tailored to the specific characteristics of each region. These measures not only aim to curb the rising trend of fall-related deaths among older adults but also have the potential to significantly reduce the economic and social impact of these events in the future, alleviating the burden on society.

## Author contributions

JMNS: conceptualization, methodology, project administration, data curation, formal analysis, writing – original draft, writing – review & editing and, final approval of the version to be submitted. RCLI: formal analysis, supervision, writing – review & editing and, final approval of the version to be submitted.

## Supplementary data

Supplementary data for this article can be found in the appendix.

## Ethical statement

This study utilizes publicly accessible secondary data; therefore, local ethics review board approval was not required

## Supplementary Material

**Figure S1.**
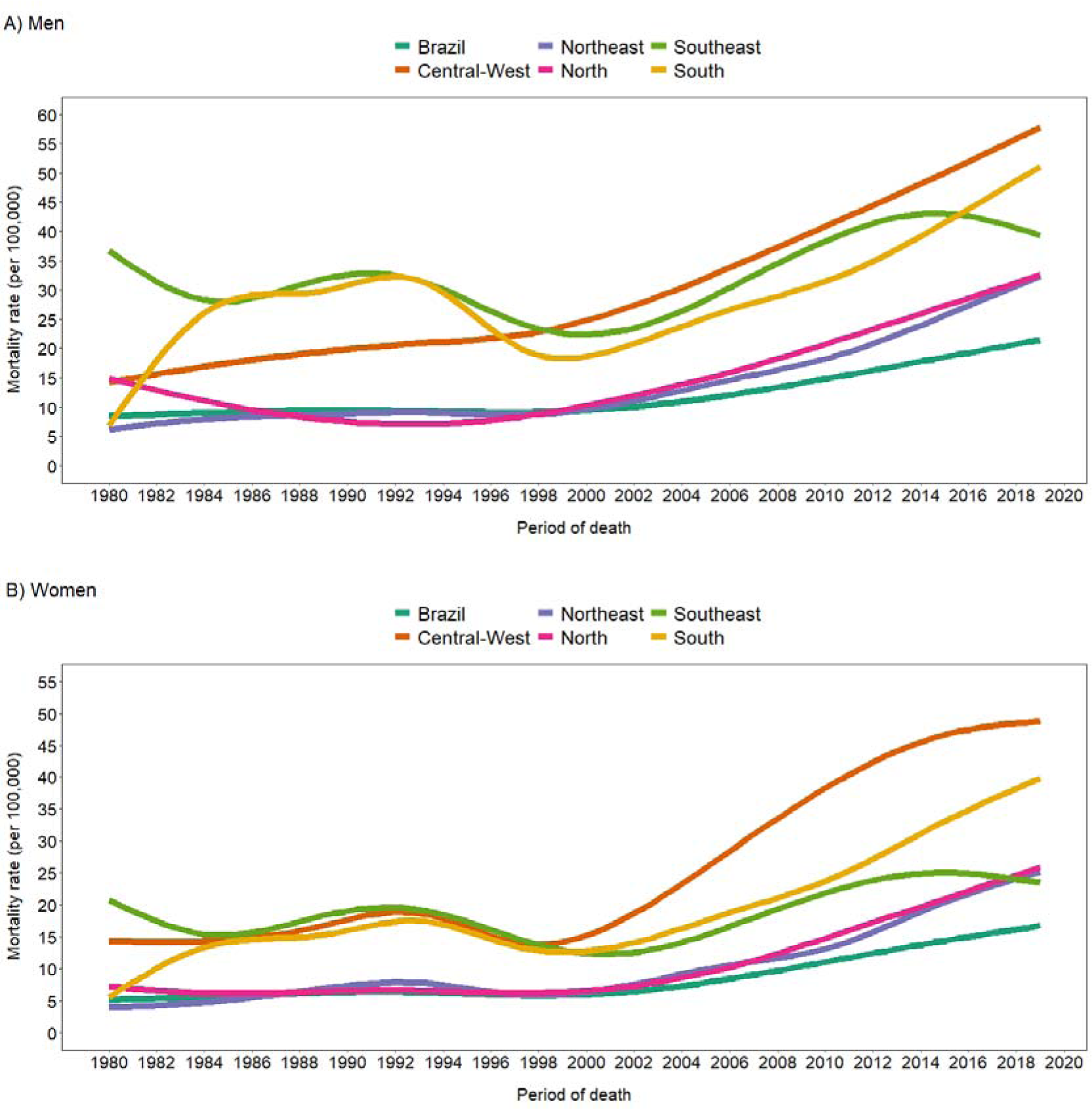
Trend in standardized fall-related mortality rates among older adults, by sex and geographic region, smoothed using third-order penalized cubic regression splines, Brazil, 1980-2019.

**Figure S2.**
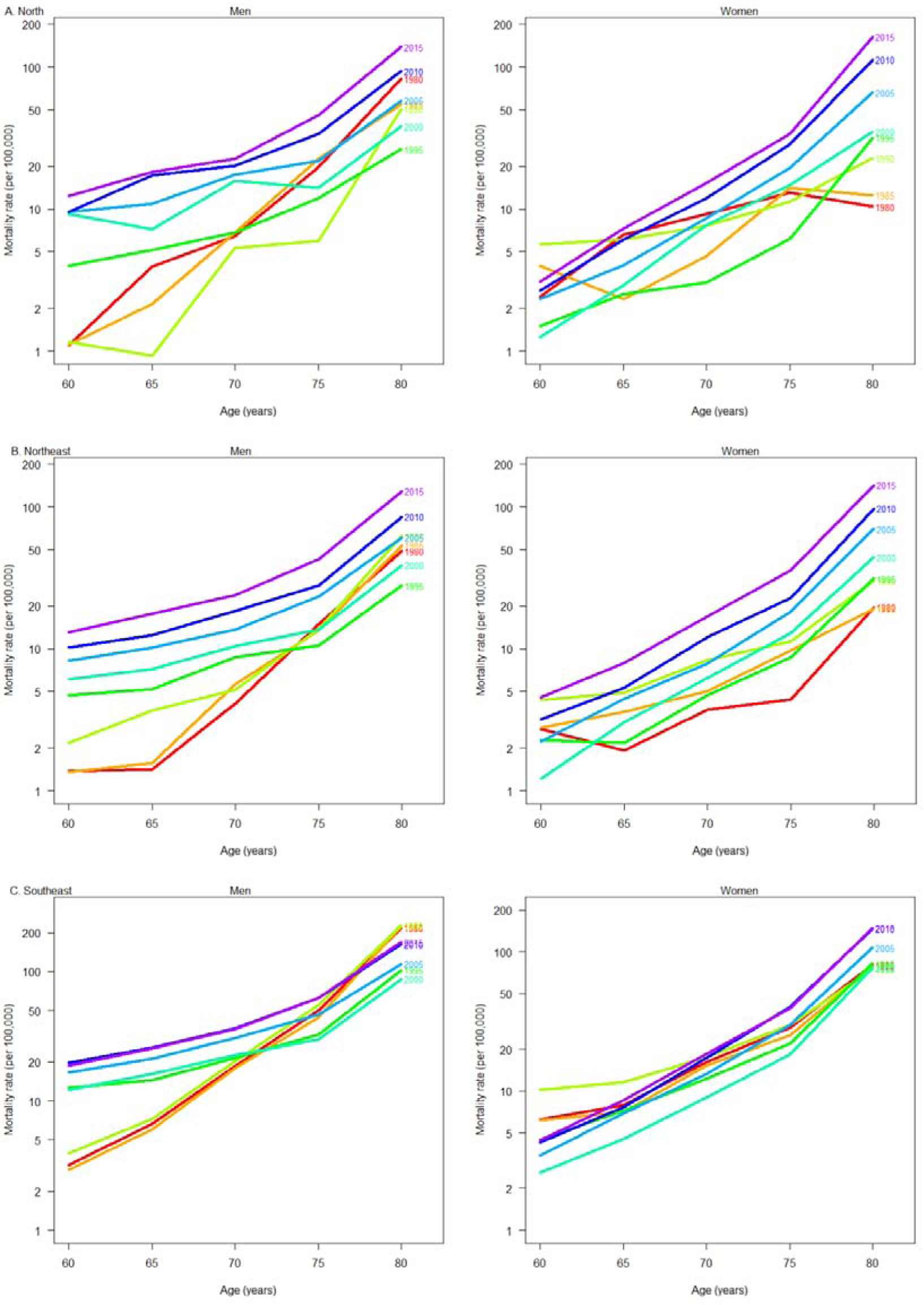
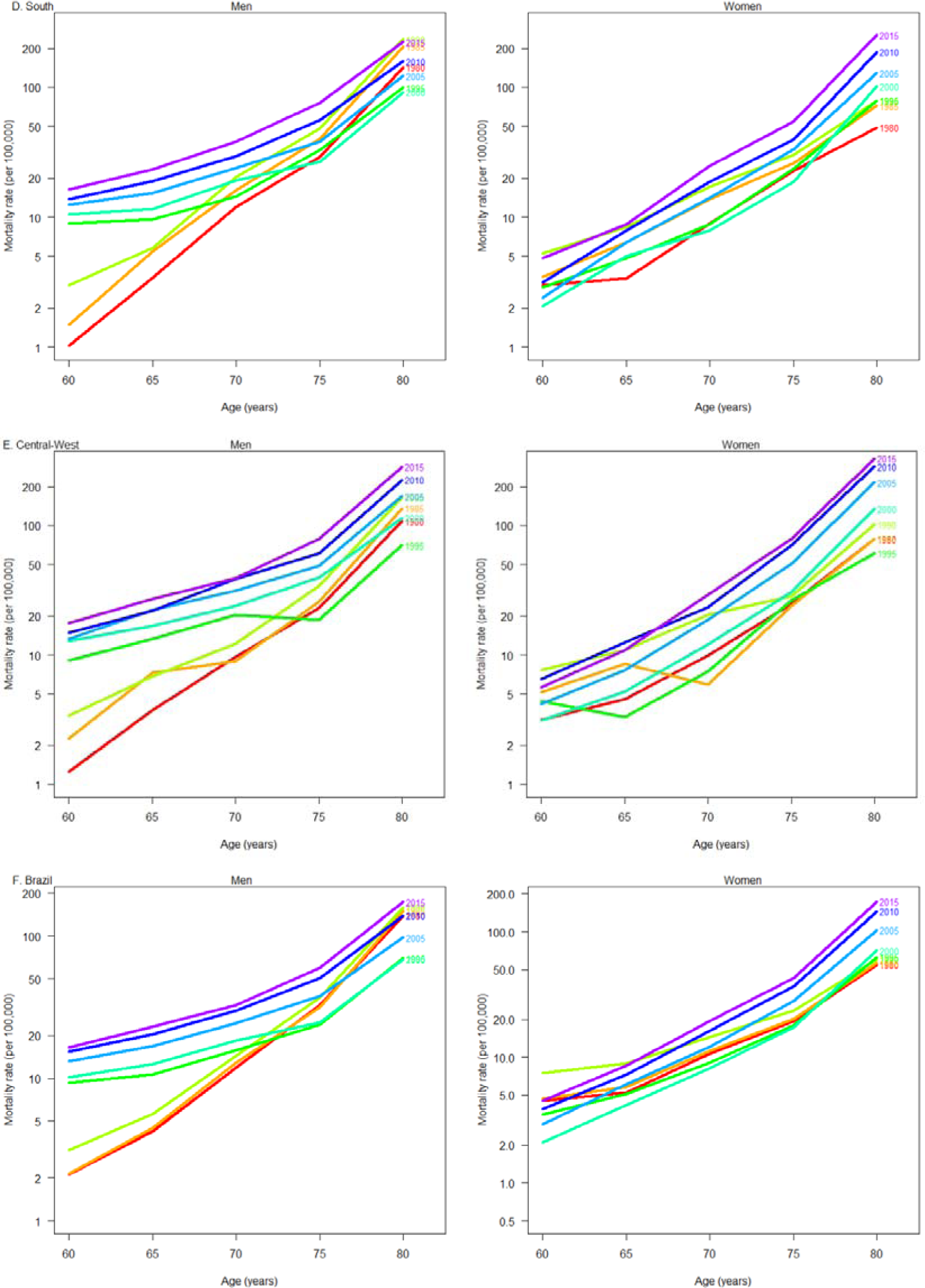
Fall-related mortality rates among older adults by age group, connected within each period, by sex and geographic region. Brazil, 1980-2019.

**Figure S3.**
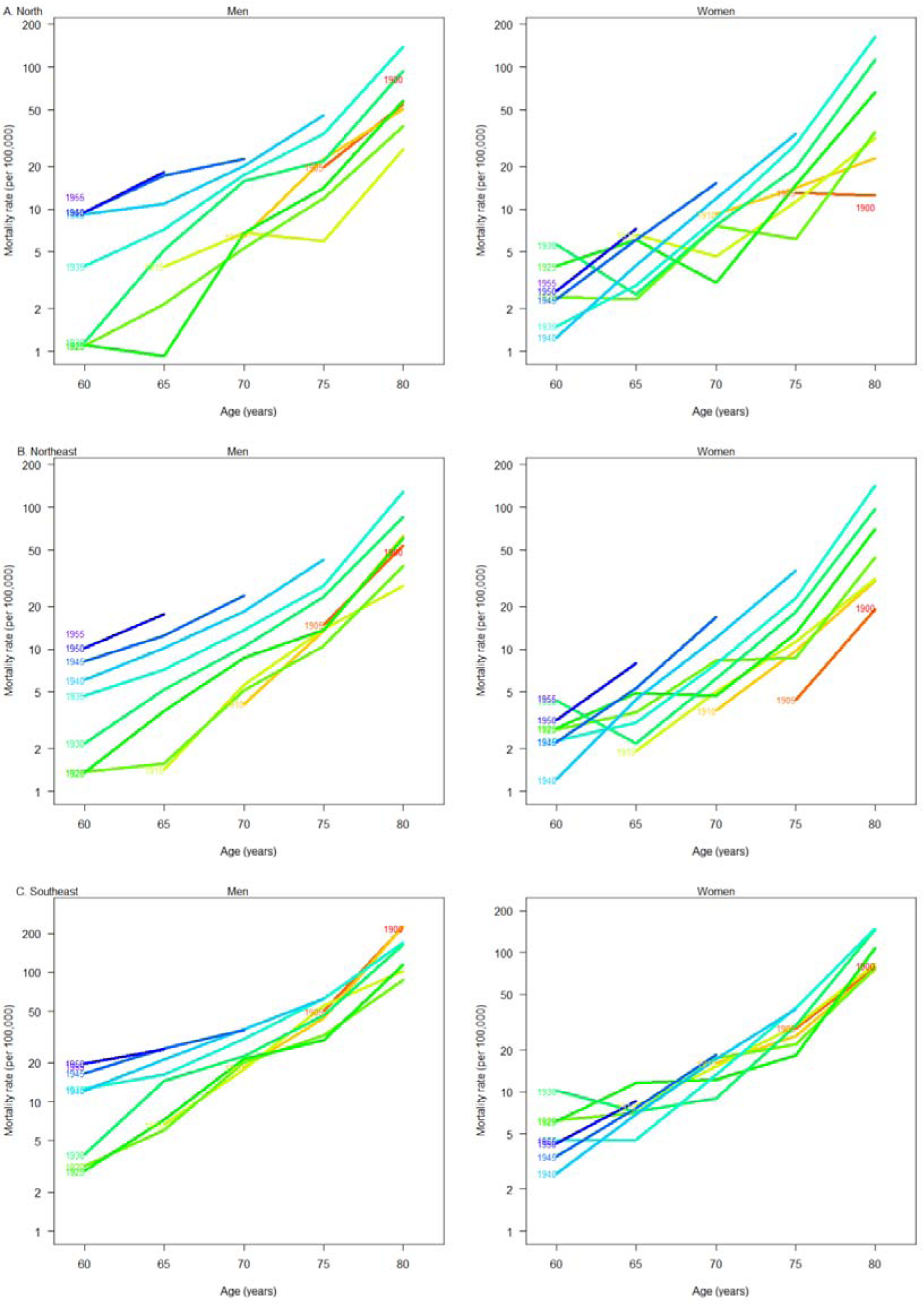
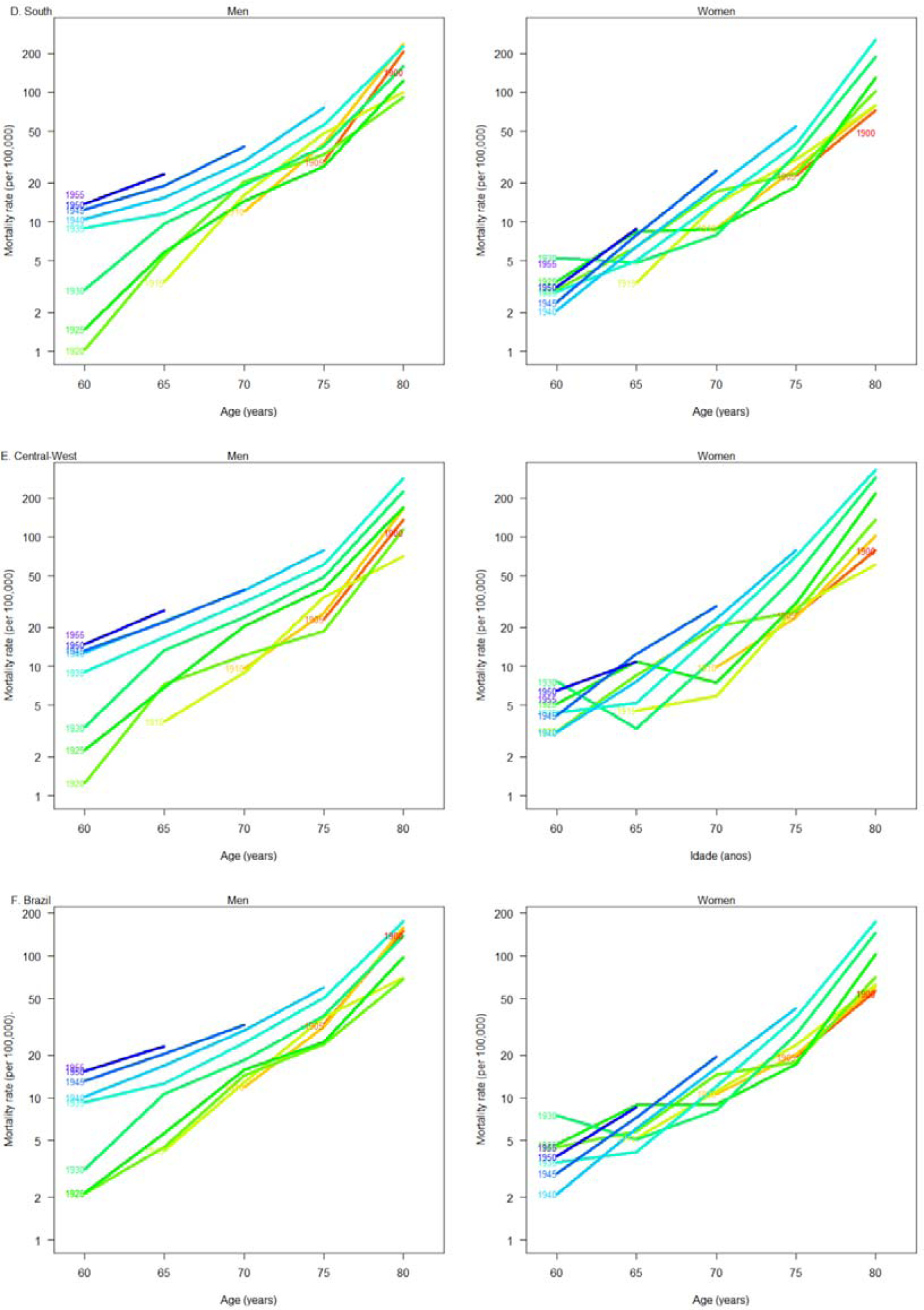
Fall-related mortality rates among older adults by age group, connected within each birth cohort, by sex and geographic region, Brazil, 1980-2019.

**Table S1.** Estimates of the age-period-cohort model adjusted for fall-related mortality rates among older men by age group, including relative risk for period and birth cohort, by geographic region, Brazil, 1980-2019.

| Men | North | Northeast | Southeast | South | Central-West | Brazil |
| --- | --- | --- | --- | --- | --- | --- |
| **Age groups (years)** |  |  |  |  |  |  |
| **Mortality rate (95% CI)** |  |  |  |  |  |  |
| 60-64 | 5.8 (4.7-7.19) | 4.8 (4.3-5.3) | 8.1 (7.7-8.5) | 5.9 (5.3-96.5) | 9.7 (8.4-11.3) | 6.9 (6.6-7.2) |
| 65-69 | 8.3 (6.8-10.05) | 6.3 (5.7-6.9) | 12.5 (11.9-13.1) | 8.9 (8.1-9.8) | 15.7 (13.8-18.0) | 10.2 (9.8-10.6) |
| 70-74 | 12.0 (9.9-14.51) | 9.9 (9.0-10.9) | 21.7 (20.6-22.8) | 17.1 (15.6-18.7) | 24.9 (21.8-28.4) | 17.32 (16.7-18.0) |
| 75-79 | 21.4 (17.6-25.97) | 17.7 (16.2-19.4) | 40.3 (38.4-42.3) | 35.0 (32.1-38.3) | 43.9 (38.6- 50.0) | 32.2 (31.1-33.4) |
| ≥80 | 68.7 (57.6-81.79) | 55.4 (51.1-60.0) | 121.4 (116.3-126.7) | 119.4 (110.4-129.2) | 171.4 (152.6-192.4) | 99.8 (96.7- 103.1) |
| **Period (95% CI)** |  |  |  |  |  |  |
| 1980-1984 | 0.42 (0.30-0.61) | 0.35 (0.30-0.41) | 0.48 (0.44-0.51) | 0.34 (0.29-0.40) | 0.28 (0.21-0.38) | 0.42 (0.40-0.45) |
| 1985-1989 | 0.48 (0.35-0.66) | 0.48 (0.42-0.54) | 0.67 (0.63-0.71) | 0.68 (0.61-0.77) | 0.45 (0.36-0.56) | 0.62 (0.59-0.65) |
| 1990-1994 | 0.36 (0.27-0.50) | 0.75 (0.68-0.84) | 1.04 (0.98-1.10) | 1.10 (1.00-1.22) | 0.67 (0.56-0.81) | 0.95 (0.91-0.99) |
| 1995-1999 | 0.63 (0.49-0.80) | 0.79 (0.71-0.88) | 1.04 (0.99-1.09) | 0.97 (0.88-1.06) | 0.73 (0.62-0.85) | 0.94 (0.90-0.98) |
| 2000-2004 |  |  |  | Reference |  |  |
| 2005-2009 | 1.15 (0.97-1.37) | 1.30 (1.20-1.41) | 1.21 (1.16-1.26) | 1.21 (1.11-1.31) | 1.15 (1.03-1.29) | 1.23 (1.19-1.27) |
| 2010-2014 | 1.37 (1.16-1.62) | 1.51 (1.40-1.64) | 1.30 (1.24-1.35) | 1.29 (1.19-1.39) | 1.30 (1.16-1.45) | 1.36 (1.32-1.40) |
| 2015-2019 | 1.66 (1.39-1.97) | 2.01 (1.86-2.17) | 1.10 (1.05-1.15) | 1.45 (1.34-1.56) | 1.48 (1.32-1.66) | 1.39 (1.34-1.43) |
| **Birth cohort (95% CI)** |  |  |  |  |  |  |
| 1900-1904 | 2.83 (1.86-4.32) | 2.50 (2.09-3.00) | 3.76 (3.46-4.08) | 3.51 (2.94-4.18) | 2.24 (1.57-3.18) | 3.29 (3.08-3.52) |
| 1905-1909 | 1.77 (1.29-2.42) | 2.10 (1.85-2.39) | 2.73 (2.57-2.90) | 2.52 (2.25-2.82) | 1.78 (1.42-2.24) | 2.42 (2.31-2.53) |
| 1910-1914 | 1.95 (1.48-2.56) | 1.49 (1.34-1.65) | 1.78 (1.70-1.88) | 1.79 (1.63-1.96) | 1.41 (1.18-1.67) | 1.65 (1.59-1.72) |
| 1915-1919 | 0.74 (0.56-0.97) | 0.76 (0.68-0.84) | 0.99 (0.94-1.04) | 1.02 (0.93-1.12) | 0.70 (0.59-0.83) | 0.90 (0.86-0.93) |
| 1920-1924 | 0.62 (0.50-0.77) | 0.70 (0.64-0.77) | 0.76 (0.72-0.80) | 0.85 (0.78-0.93) | 0.68 (0.59-0.78) | 0.73 (0.70-0.76) |
| 1925-1929 | 0.71 (0.59-0.85) | 0.84 (0.78-0.91) | 0.77 (0.73-0.80) | 0.80 (0.74-0.87) | 0.86 (0.77-0.97) | 0.78 (0.76-0.81) |
| 1930-1934 |  |  |  | Reference |  |  |
| 1935-1949 | 1.19 (1.04-1.36) | 1.13 (1.06-1.21) | 1.26 (1.22-1.30) | 1.29 (1.21-1.37) | 1.12 (1.02-1.22) | 1.23 (1.20-1.26) |
| 1940-1944 | 1.29 (1.10-1.51) | 1.22 (1.14-1.31) | 1.39 (1.34-1.44) | 1.47 (1.37-1.58) | 1.23 (1.10-1.36) | 1.33 (1.30-1.37) |
| 1945-1949 | 1.32 (1.11-1.58) | 1.25 (1.16-1.36) | 1.58 (1.52-1.65) | 1.62 (1.49-1.75) | 1.09 (0.97-1.23) | 1.44 (1.39-1.49) |
| 1950-1954 | 1.28 (1.04-1.58) | 1.40 (1.28-1.54) | 1.87 (1.78-1.97) | 1.81 (1.65-1.98) | 1.17 (1.01-1.35) | 1.64 (1.58-1.70) |
| 1955-1959 | 1.28 (0.97-1.69) | 1.36 (1.20-1.54) | 2.11 (1.97-2.26) | 1.92 (1.70-2.18) | 1.22 (1.01-1.49) | 1.73 (1.65-1.82) |

**Table S2.** Estimates of the age-period-cohort model adjusted for fall-related mortality rates among older women by age group, including relative risk for period and birth cohort, by geographic region, Brazil, 1980-2019.

| Women | North | Northeast | Southeast | South | Central-West | Brazil |
| --- | --- | --- | --- | --- | --- | --- |
| **Age groups (years)** |  |  |  |  |  |  |
| **Mortality rate (95% CI)** |  |  |  |  |  |  |
| 60-64 | 3.1 (2.3-4.2) | 2.5 (2.2-2.8) | 3.8 (3.6-4.1) | 2.6 (2.3-3.0) | 4.0 (3.3-4.8) | 3.3 (3.1-3.4) |
| 65-69 | 4.5 (3.4-5.8) | 3.4 (3.0-3.8) | 5.5 (5.2-5.9) | 4.8 (4.3-5.4) | 5.8 (4.8-6.9) | 4.8 (4.6-5.1) |
| 70-74 | 7.4 (5.8-9.4) | 6.3 (5.6-6.9) | 10.6 (10.0-11.2) | 10.0 (9.0-11.1) | 11.3 (9.5-13.3) | 9.3 (8.9-9.7) |
| 75-79 | 14.0 (11.1-17.6) | 11.5 (10.5-12.7) | 20.9 (19.8-22.1) | 21.2 (19.3-23.3) | 29.8 (25.6-34.6) | 18.7 (17.9-19.4) |
| ≥80 | 45.6 (36.9-56.3) | 41.7 (38.3-45.4) | 73.6 (70.1-77.2) | 86.8 (79.8-94.4) | 112.0 (97.9-128.2) | 67.4 (65.1-69.9) |
| **Period (95% CI)** |  |  |  |  |  |  |
| 1980-1984 | 1.86 (1.33-2.60) | 0.90 (0.77-1.06) | 2.01 (1.87-2.16) | 1.28 (1.10-1.48) | 1.11 (0.84-1.47) | 1.61 (1.52-1.70) |
| 1985-1989 | 1.03 (0.74-1.43) | 1.10 (0.97-1.24) | 1.66 (1.56-1.77) | 1.44 (1.28-1.62) | 1.12 (0.89-1.40) | 1.46 (1.39-1.53) |
| 1990-1994 | 1.15 (0.87-1.53) | 1.28 (1.15-1.42) | 1.86 (1.76-1.97) | 1.56 (1.40-1.73) | 1.55 (1.29-1.85) | 1.65 (1.58-1.72) |
| 1995-1999 | 0.68 (0.50-0.91) | 0.83 (0.74-0.92) | 1.28 (1.21-1.35) | 1.02 (0.92-1.13) | 0.79 (0.65-0.95) | 1.09 (1.04-1.13) |
| 2000-2004 |  |  |  | Reference |  |  |
| 2005-2009 | 1.42 (1.14-1.77) | 1.56 (1.43-1.70) | 1.47 (1.40-1.54) | 1.56 (1.42-1.70) | 1.70 (1.48-1.95) | 1.51 (1.46-1.57) |
| 2010-2014 | 2.50 (2.03-3.07) | 2.35 (2.16-2.55) | 2.02 (1.92-2.11) | 2.16 (1.99-2.35) | 2.58 (2.26-2.95) | 2.17 (2.09-2.24) |
| 2015-2019 | 4.27 (3.47-5.27) | 4.06 (3.74-4.40) | 2.32 (2.22-2.44) | 3.22 (2.97-3.49) | 3.19 (2.79-3.64) | 2.94 (2.84-3.04) |
| **Birth cohort (95% CI)** |  |  |  |  |  |  |
| 1900-1904 | 0.12 (0.06-0.25) | 0.52 (0.42-0.65) | 0.54 (0.50-0.59) | 0.44 (0.36-0.54) | 0.63 (0.44-0.90) | 0.50 (0.47-0.54) |
| 1905-1909 | 0.34 (0.23-0.52) | 0.42 (0.36-0.49) | 0.65 (0.61-0.69) | 0.62 (0.55-0.71) | 0.66 (0.52-0.84) | 0.59 (0.56-0.62) |
| 1910-1914 | 0.56 (0.42-0.75) | 0.61 (0.54-0.68) | 0.60 (0.57-0.64) | 0.64 (0.57-0.71) | 0.62 (0.52-0.75) | 0.59 (0.57-0.62) |
| 1915-1919 | 0.87 (0.67-1.13) | 0.83 (0.75-0.92) | 0.84 (0.79-0.88) | 0.89 (0.81-0.98) | 0.66 (0.54-0.79) | 0.82 (0.79-0.85) |
| 1920-1924 | 0.72 (0.57-0.92) | 1.04 (0.95-1.14) | 0.95 (0.90-1.00) | 1.13 (1.04-1.23) | 1.19 (1.03-1.37) | 0.99 (0.95-1.02) |
| 1925-1929 | 1.02 (0.85-1.23) | 1.07 (0.99-1.16) | 0.98 (0.94-1.02) | 0.95 (0.88-1.03) | 1.12 (1.00-1.25) | 1.00 (0.97-1.03) |
| 1930-1934 |  |  |  | Reference |  |  |
| 1935-1949 | 0.83 (0.72-0.96) | 0.84 (0.79-0.90) | 0.88 (0.85-0.91) | 0.91 (0.85-0.97) | 0.93 (0.85-1.02) | 0.88 (0.86-0.90) |
| 1940-1944 | 0.58 (0.48-0.71) | 0.77 (0.71-0.83) | 0.81 (0.77-0.84) | 0.82 (0.76-0.89) | 0.82 (0.73-0.93) | 0.79 (0.76-0.81) |
| 1945-1949 | 0.50 (0.40-0.63) | 0.65 (0.59-0.71) | 0.71 (0.67-0.75) | 0.75 (0.68-0.82) | 0.79 (0.69-0.91) | 0.70 (0.67-0.72) |
| 1950-1954 | 0.37 (0.28-0.48) | 0.56 (0.50-0.63) | 0.62 (0.58-0.67) | 0.57 (0.50-0.64) | 0.60 (0.51-0.72) | 0.58 (0.56-0.61) |
| 1955-1959 | 0.23 (0.16-0.34) | 0.44 (0.38-0.52) | 0.50 (0.45-0.55) | 0.58 (0.48-0.69) | 0.44 (0.34-0.57) | 0.47 (0.44-0.51) |

